# Symptoms of COVID-19 in a population-based cohort study

**DOI:** 10.1101/2021.03.20.21254040

**Authors:** Sana M. Khan, Leslie V. Farland, Erika Austhof, Melanie L. Bell, Collin J. Catalfamo, Zhao Chen, Felina Cordova-Marks, Kacey C. Ernst, Pamela Garcia-Filion, Kelly M. Heslin, Joshua Hoskinson, Megan L Jehn, Emily C.S. Joseph, Connor P. Kelley, Yann Klimentidis, Stephanie Russo Carroll, Lindsay N. Kohler, Kristen Pogreba-Brown, Elizabeth T. Jacobs

## Abstract

Accurate diagnosis of potential SARS-CoV-2 infections by symptoms is one strategy for continuing global surveillance, particularly in low-resource communities. We conducted a prospective, population-based cohort study, the Arizona CoVHORT, among Arizona residents to elucidate the symptom profile of laboratory-confirmed COVID-19 participants(16.2%) compared to laboratory-confirmed negative(22.4%) and untested general population participants(61.4%). Among the 1514 study participants, those who were COVID-19 positive were more likely to be Hispanic(33.5%) and more likely to report obesity ≥ 30 kg/m^2^(34.7%) compared to COVID-19 negative participants(19.2%; 31.0%) and untested CoVHORT participants(13.8%; 23.8%). Of the 245 laboratory-confirmed COVID-19 cases, 15.0% reported having had no symptoms. Of those that did report symptoms, the most commonly-reported first symptoms were sore throat(19.0%), headache(15.5%), cough(12.7%), runny nose/cold-like symptoms(12.1%), and fatigue(12.0%). In adjusted logistic regression models, COVID-19 positive participants were more likely than negative participants to experience loss of taste and smell(OR:35.7; 95% CI 18.4-69.5); bone or nerve pain(OR:17.9; 95% CI 6.7-47.4), vomiting(OR:10.8; 95% CI 3.1-37.5), nausea(OR:10.5; 95% CI 5.5-19.9), and headache(OR:8.4; 95% CI 5.6-12.8). When comparing confirmed COVID-19 cases with confirmed negative or untested participants, the pattern of symptoms that discriminates SARS-CoV-2 infection from those arising from other potential circulating pathogens may differ from general reports of symptoms among cases alone.

## Introduction

In late 2019, the novel coronavirus SARS-CoV-2 was first recognized in China among patients who presented with pneumonia and the first scientific report appeared shortly thereafter (1). On March 11^th^, 2020 the World Health Organization declared COVID-19 a pandemic. The pathogen has had multiple impacts on individual and societal wellbeing arising from both biological effects of the virus and policy-based mitigation. The majority of those infected with acute COVID-19 will go on to recover, though approximately 10-20% of COVID-19 patients overall will develop a severe case of disease, and may suffer from stroke, pneumonia, or acute respiratory distress syndrome (ARDS) and require intensive care and ventilation (2, 3).

Individuals are likely be most infectious during the early phases of the disease, when symptoms may be comparatively mild; therefore, it is important to elucidate the reported symptom patterns of COVID-19 patients compared to both laboratory-confirmed negative individuals and population-based controls. Several risk factors have been associated with disease susceptibility and severity including increasing age (4), male sex (2, 5, 6), and current or former smoking (3), which may also affect symptomology. Further, important differences in disease incidence and severity by race and ethnicity have emerged, with Native Americans, African-Americans, and Latinos having higher COVID-19 prevalence, hospitalization, and mortality rates compared to non-Hispanic whites (7). It is presently not known if reports of symptoms or symptom patterns vary by these factors as well.

A recent meta-analysis of over 24,000 patients across nine countries reported on COVID-19 symptom presentation. In this work, the most commonly-reported symptoms among people with COVID-19 were fever (78% of COVID-19 patients reporting), cough (57%), and fatigue (31%) (8). In comparison, another study conducted among European patients (n=1420) with mild or moderate COVID-19 found that the most frequently reported symptoms were headache (70%), loss of smell (70%), and obstruction of the nasal passages (68%) (9). The authors of another study, the objective of which was to develop a better symptom modeling algorithm to aid targeted testing, concluded that fever and cough should be used as the key symptoms for rapid COVID-19 screening given their high sensitivity (10). However, a major limitation of studies conducted to date is the lack of comparison of patient-reported symptoms to those of uninfected individuals. To our knowledge, no prior research has compared the prevalence of non-specific symptoms such as headache, fever, and runny nose between confirmed COVID-19-positive cases, confirmed COVID-19-negative cases, and population-based comparison groups.

Since COVID-19 community transmission began, Arizona has twice experienced severe COVID-19 surges, with more than 850,000 infections and 16,000 COVID-19-related deaths as of March, 2021. To address this epidemiological challenge, in May 2020, we initiated a large, prospective, population-based cohort in Arizona of racially- and ethnically-diverse residents in order to rigorously investigate factors contributing to variability in natural COVID-19 disease history including incidence, progression, resolution, and chronic outcomes of infection (11). This COVID-19 cohort, dubbed The Arizona CoVHORT, provides a rich data source for multiple areas of inquiry related to the pandemic. The objective of the present work was to determine which symptoms were reported with the greatest frequency among participants who tested positive for COVID-19 as compared to participants who tested negative for COVID-19 and untested participants, while controlling for potential confounders such as age, ethnicity, and sex. The findings of this paper will aid in the identification of symptoms that differentiate COVID-19 from other circulating infections or conditions, such as allergies.

## Materials and Methods

### Study Participants

The overall goal of the CoVHORT is to continuously enroll Arizonans into a cohort study to track both the acute and long-term phases of infection with SARS-CoV-2. The present analysis includes data through October 31^st^, 2020, five months since the cohort was launched on May 28^th^, 2020. Several recruitment methods were employed, which have been described in detail previously (11). Briefly, the primary sources of recruitment have been through case investigations in a partnership with the Arizona Department of Health Services and the COVID-19 Antibody Testing Initiative (CATI), both of which have allowed for inclusion of laboratory-confirmed COVID-19 positive participants. By October 31^st^, 2020, a total of 176 COVID-19-positive participants had been recruited through health department case investigations and 10 through our partnership with CATI; further, a total of 168 participants who had COVID-19-negative results were recruited via CATI.

A comprehensive mailing list was purchased that provides information on 2.2 million residents in Arizona. To recruit the population-based comparison group, a total of 17,500 postcards were mailed to a simple random sample of Pima, Yuma, and Pinal counties beginning in July 2020. Consistent with the Dillman method to maximize participation and minimize bias (12), three phased mailings of recruitment postcards occurred every two weeks. Participant-provided information from baseline surveys was used to exclude those who had already enrolled from subsequent phases of the mailing campaign. Each list was screened prior to each mailing to reduce the number of undeliverable postcards. In total, we have contacted more than 51,000 residences from these phased mailing campaigns. Method of recruitment is recorded for all participants allowing sensitivity analyses to be conducted within subgroups.

### Survey Instruments

All participants included in the CoVHORT were sent identical structured electronic questionnaires at baseline, regardless of COVID-19 status. Participants were asked if they had tested for the virus that causes COVID-19 with a nasal swab, throat swab, or saliva test since January 2020. Participants were classified as untested, positive or negative based on their results. Participants, regardless of COVID-19 test status, were asked, “Since January 1, have you experienced a sudden illness that led you to believe you had COVID19?” If they answered “yes”, all participants, regardless of case status, were asked to indicate which symptoms they had experienced since January 2020 from a list based upon prior reports in the literature, as well as through an open-text field. Further, COVID-19 positive participants were queried regarding the first symptom that they recalled having experienced. Information regarding health and medical history was collected, along with other demographic data, including age, sex, race, and ethnicity, as well as for weight, height, and smoking status.

### Statistical analysis

Data were analyzed to describe the COVID-19 symptom profile, estimate the prevalence of individual symptoms, and identify differences between COVID-19-positive, COVID-19-negative, and untested participants. Individual variables were summarized and reported using appropriate statistical measures: mean [standard deviation (SD)] for continuous and percent (%) for categorical variables. We compared the participant characteristics at baseline and number of symptoms (0 symptoms, 1-6 symptoms, 7-9 symptoms, 10-16 symptoms) using ordered logistic regression. Nonparametric analogs were used when appropriate. Additionally, a logistic regression model was fit for each symptom to measure the association with COVID-19-positive status. Statistical significance was defined as an alpha of 0.05, with two-sided alternative hypotheses. Data analyses was conducted using Stata 16.0 (College Station, TX).

## Results

As of October 31^st^, 2020, the CoVHORT study had enrolled a total of 1,514 participants, 245 (16.2%) of whom had lab-confirmed COVID-19. Of the remaining 1,269 participants, 339 (22.4%) participants had tested negative for COVID-19 and 930 (61.4%) had not been tested (Table 1). The participants were majority female (63.0%) and white (86.8%) and had a mean (SD) age of 47.8 (16.8) years. COVID-19-positive participants were younger (39.2 years) than COVID-19-negative participants (47.5 years), and participants who had not been tested for COVID-19 (50.1 years). COVID-19 positive participants were more likely to be Hispanic (33.5%), compared to COVID-19-negative participants (19.2%) and untested CoVHORT participants (13.8%). COVID-19-positive participants were more likely to have a body mass index (BMI) of greater than 30 kg/m^2^ (34.7%) compared with COVID-19-negative participants (31.0%) and untested CoVHORT participants (23.8%). In addition, those with COVID-19 were more likely to be never smokers (93.5 vs. 86.1 and 90.3% for negative and untested participants, respectively).

**Table 1.** Demographic characteristics of CoVHORT participants who were laboratory-confirmed positive for COVID-19, those who were tested and were negative for COVID-19, and those without COVID-19 test results in the CoVHORT population.

| Characteristics at study entry (baseline) | Lab-confirmed COVID-19 status |  |  |
| --- | --- | --- | --- |
|  | Untested participants<sup>1</sup><br>n=930 | COVID-19 negative<sup>2</sup><br>n=339 | COVID-19 positive<sup>3</sup><br>n=245 |
| Age (years, mean ± sd) | 50.1 (16.5) | 47.5 (15.9) | 39.2 (16.8) |
| Age (median, IQR) | 51 (36,63) | 48 (34,61) | 38 (24, 51) |
| Age (range) | 12-96 | 18-86 | 12-80 |
| Sex (%)<sup>4</sup> |  |  |  |
| Male | 319 (34.3) | 143 (42.2) | 89 (36.3) |
| Female | 605 (65.0) | 193 (56.9) | 155 (63.3) |
| Non-binary | 6 (0.7) | 2 (0.6) | 1 (0.4) |
| Ethnicity (n, %)<sup>5</sup> |  |  |  |
| Hispanic | 128 (13.8) | 65 (19.2) | 82 (33.5) |
| Non-Hispanic | 751 (80.8) | 255 (75.2) | 150 (61.2) |
| BMI (kg/m<sup>2</sup>) |  |  |  |
| < 18.5 | 8 (0.9) | 6 (1.8) | 6 (2.5) |
| 18.5 – 24.9 | 379 (40.8) | 117 (34.5) | 88 (35.9) |
| 25.0 – 29.9 | 306 (32.9) | 105 (31.0) | 64 (26.1) |
| 30.0 – 39.9 | 188 (20.2) | 80 (23.6) | 73 (29.8) |
| ≥ 40 | 33 (3.6) | 25 (7.40) | 12 (4.9) |
| Smoking status (n, %) |  |  |  |
| Never | 840 (90.3) | 292 (86.1) | 229 (93.5) |
| Occasionally | 30 (3.2) | 24 (7.1) | 13 (5.3) |
| Regularly | 26 (2.8) | 13 (3.8) | 1 (0.41) |

<sup>1</sup>All participants in CoVHORT who do not have a laboratory-confirmed result; <sup>2</sup>PCR or antibody negative; <sup>3</sup>PCR-positive; <sup>4</sup>Prefer not to answer (n=1); <sup>5</sup>Missing data for ethnicity (n=83); BMI (n=24); smoking (n=46);

Of the 245 lab-confirmed COVID-19-positive participants, the majority (85.0%) reported having experienced at least one symptom at baseline, while the remaining 38 participants (15.0%) were asymptomatic, having reported never experiencing any symptoms (Table 2). When asked to self-rate the severity of their illness on a scale of 0-10, those who reported 10-16 symptoms reported a mean (SD) severity score of 6.5 (2.0), while participants with 7-9 symptoms reported a mean severity score of 5.5 (2.4), and participants with 1-6 symptoms reported a mean severity score of 3.3 (2.1).

**Table 2.**
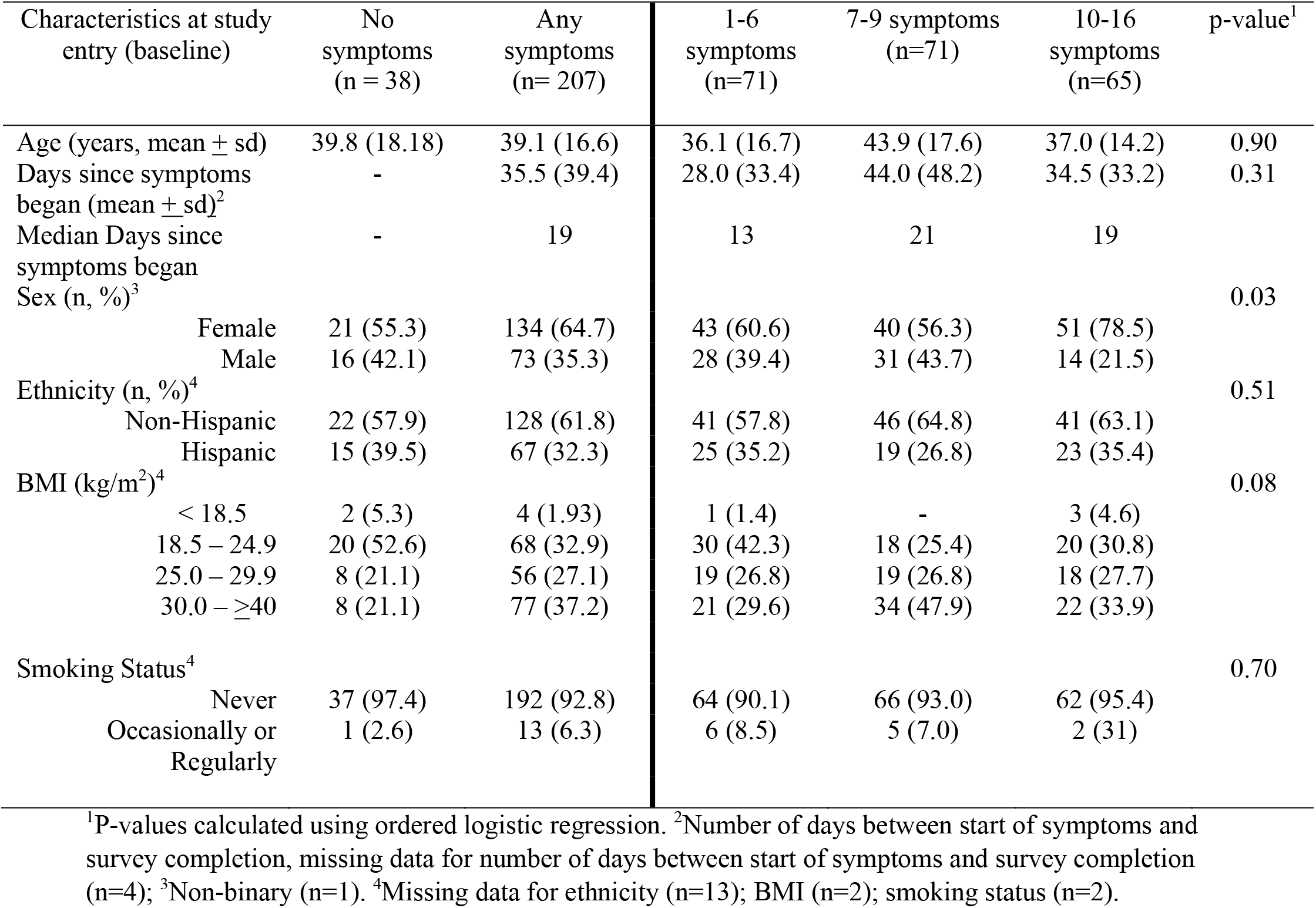
Characteristics of COVID-19 positive study participants (n=245) by reported number of COVID-19 disease symptoms.

Figure 1 displays the first symptom reported by COVID-19-positive study subjects who stated that they had experienced at least one symptom. The most common first symptoms were sore throat (19.0%), headache (15.5%), cough (12.7%), runny nose/cold-like symptoms (12.1%), and fatigue (12.0%). As shown in Table 3, other common symptoms that lab-confirmed COVID-19-positive participants reported at any time in their disease course included fatigue (71.8%), headache (64.5%), loss of taste or smell (57.1%), aches and pains or sore muscles (58.0%), and cough (54.3%). COVID-19-positive participants had greater odds of reporting fever, sore throat, difficulty breathing or shortness of breath, chills, diarrhea, headache, and “other symptoms” when compared to participants who tested negative for COVID-19 and participants who were never tested for COVID-19. While the magnitude of effect for these latter symptoms was smaller, all results were statistically significant. No differences between groups were observed for rash on skin, discoloration of fingers or toes, and conjunctivitis.

**Figure 1.**
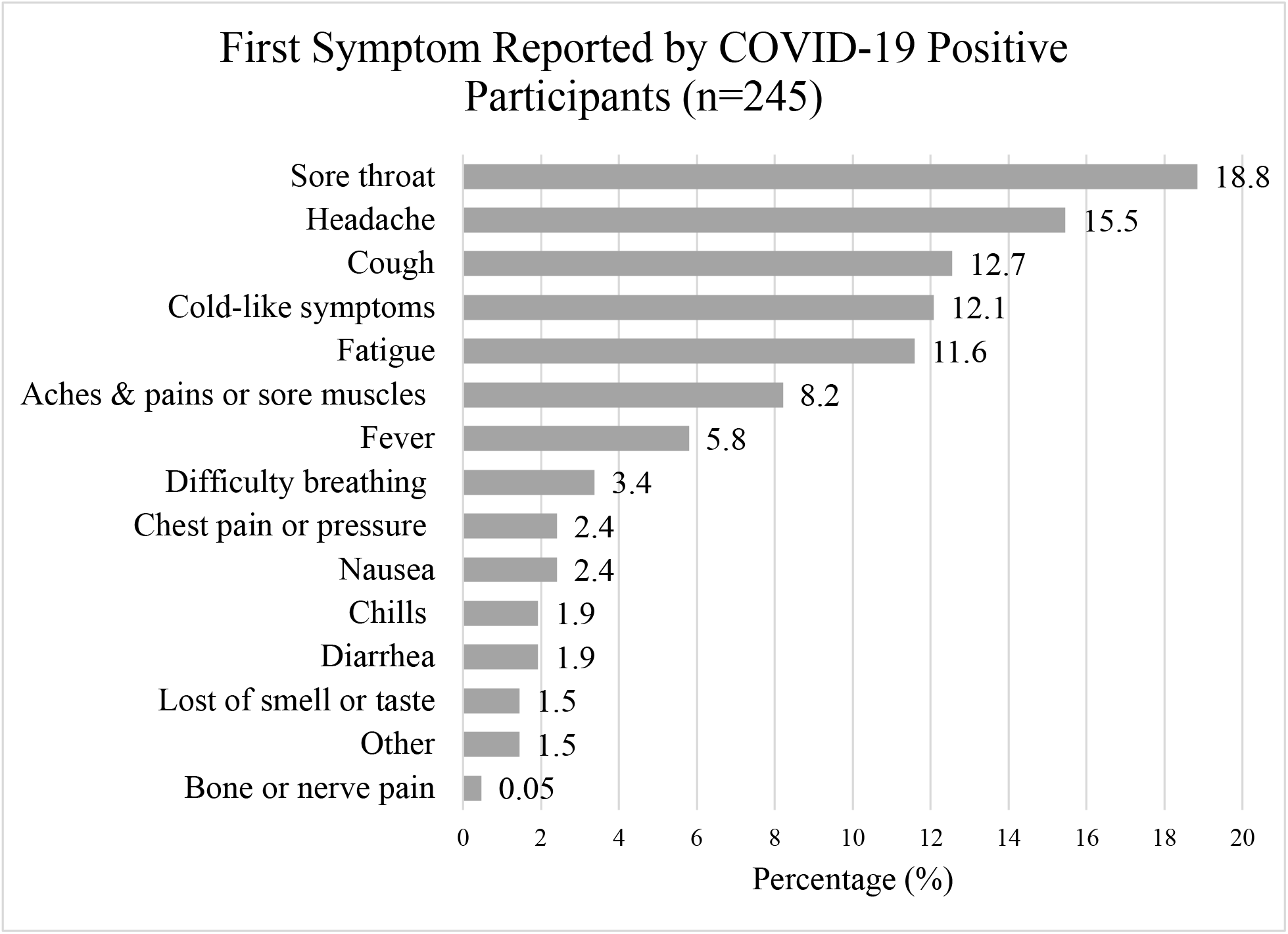
First symptom reported by participants who are laboratory-confirmed positive COVID-19 cases.

**Table 3.** Symptom characteristics of CoVHORT participants by case status, adjusted by age, sex, and ethnicity.

| Reported symptoms<br>at study entry<br>(baseline) | COVID-19<br>positive<sup>1</sup><br>n=245 | Untested<br>participants<sup>2</sup><br>n=930 | COVID-19<br>negative<sup>3</sup><br>n=339 | Positive vs<br>Untested | Positive vs<br>Negative |
| --- | --- | --- | --- | --- | --- |
|  | n (%) | n (%) | n (%) | OR (95% C I) | OR (95% CI) |
| Fatigue | 176 (71.8) | 170 (18.3) | 87 (25.7) | 11.4 (8.0, 16.3) | 8.3 (5.5, 12.5) |
| Headache | 158 (64.5) | 120 (12.9) | 60 (17.7) | 11.3 (8.0, 16.2) | 8.4 (5.6, 12.8) |
| Aches and pains or<br>sore muscles | 142 (58.0) | 130 (14.0) | 66 (19.5) | 8.5 (6.0, 11.9) | 6.4 (4.3, 9.7) |
| Loss of smell/taste | 140 (57.1) | 48 (5.2) | 12 (3.5) | 21.1 (14.0, 32.0) | 35.7 (18.4, 69.5) |
| Cough | 133 (54.3) | 156 (16.8) | 75 (22.1) | 5.5 (3.9, 7.6) | 4.0 (2.7, 5.9) |
| Fever | 122 (49.8) | 120 (12.9) | 61 (18.0) | 6.7 (4.8, 9.5) | 5.2 (3.5, 7.9) |
| Runny nose/cold-like<br>symptoms | 115 (46.9) | 105 (11.3) | 43 (12.7) | 6.4 (4.5, 9.1) | 6.3 (4.0, 9.8) |
| Chills | 104 (42.5) | 89 (9.6) | 39 (11.5) | 6.9 (4.7, 9.9) | 6.8 (4.3, 10.7) |
| Sore throat | 101 (41.2) | 123 (13.2) | 61 (18.0) | 3.9 (2.7, 5.5) | 2.9 (2.0, 4.4) |
| Difficulty breathing<br>or shortness of breath | 92 (37.6) | 94 (10.1) | 48 (14.2) | 5.4 (3.7, 7.8) | 4.2 (2.7, 6.6) |
| Diarrhea | 81 (33.0) | 50 (5.4) | 19 (5.6) | 8.4 (5.5, 12.9) | 9.0 (5.1, 16.1) |
| Nausea | 79 (32.2) | 34 (3.7) | 14 ( 4.1) | 10.9 (6.6, 17.5) | 10.5 (5.5, 19.9) |
| Chest pain or<br>pressure | 74 (30.2) | 61 (6.6) | 41 (12.1) | 6.4 (4.2, 9.7) | 3.4 (2.1, 5.3) |
| Bone pain/nerve pain | 47 (19.2) | 10 (1.1) | 5 ( 1.5) | 25.5 (12.0, 54.2) | 17.9 (6.7, 47.4) |
| Vomiting | 24 (9.8) | 9 (1.0) | 3 ( 0.9) | 8.7 (3.8, 20.4) | 10.8 (3.1, 37.5) |
| Other | 17 (6.9) | 9 (0.97) | 3 ( 0.9) | 8.4 (3.3, 21.4) | 8.1 (2.2, 29.2) |
| Rash on skin | 15 (6.1) | 8 (0.9) | 4 ( 1.2) | 10.9 (4.1, 28.7) | 7.9 (0.86, 73.4) |
| Discoloration of<br>fingers/toes | 5 (2.0) | 3 (0.3) | 1 ( 0.3) | 7.1 (1.6, 31.9) | 7.0 (0.82, 60.7) |
| Loss of speech or<br>movement | 2 (0.8) | 1 (0.1) | - | 7.0 (0.5, 91.8) | - |
| Conjunctivitis | 2 (0.8) | 7 (0.8) | 2 ( 0.6) | 1.21 (0.22, 6.6) | 1.2 (0.16, 8.6) |

<sup>1</sup>PCR-positive cases; <sup>2</sup>participants in CoVHORT who do not have a laboratory-confirmed result; <sup>3</sup>PCR or antibody negative.

After adjusting for age, ethnicity, and sex, COVID-19 positive participants were more likely than negative participants to experience loss of taste and smell (OR 35.7, 95% CI 18.5-69.5), bone or nerve pain (OR 17.9, CI 6.7-47.4), vomiting (OR 10.8, CI 3.1-37.5), nausea (OR 10.5, CI 5.5-19.9), and headache (OR 8.4, CI 5.6-12.8) (Table 3). Similarly, the symptoms with the strongest association when comparing COVID-19-positive cases with the untested participants were loss of taste or smell (OR 21.1, CI 14.0-32.0), bone/nerve pain (OR 25.5, CI 12.0-54.2.0), headache (OR 11.3, CI 8.0-16.2), nausea (OR 10.9, CI 6.6-17.5), and vomiting (OR 8.7, CI 3.8-20.4).

## Discussion

We determined that lab-confirmed COVID-19 cases differed in age, ethnicity, BMI and smoking status from COVID-negative participants, and untested cohort members. These same factors were associated with reported symptom severity. The most commonly reported first symptoms among COVID-19 positive participants were sore throat, followed by headache, cough, runny nose/cold-like symptoms, and fatigue. Discriminating symptoms for COVID-19-positivity included loss of taste and smell and bone or nerve pain.

Individuals identifying as Hispanic in CoVHORT constituted 33.5% of the recruited COVID-positive participants, mirroring the broader statewide case composition reported by the Arizona Department of Health Services(13). By comparison, they constituted far fewer of the lab-negative and untested groups. As discussed by Macias Gil et al.(14), the burden of COVID-19 on communities of color has been far more extreme due to extant healthcare disparities, with greater rates of hospitalizations and deaths among U.S. Hispanics as compared to whites being reported in other studies (14). Further, because publicly-available COVID-19 data by race or ethnicity may have missing values, it is critical to continue to follow up the health outcomes of this medically-vulnerable group.

Differences in disease outcomes by body size have been well-documented. In the first large study of COVID-19 patients in the United States, obesity was determined to be a major risk factor for hospitalization (3), but it remains unclear whether this finding is attributable to comorbidities that are themselves associated with both larger body size and with severe COVID-19. In the present work, only those with a BMI of greater than 30 kg/m^2^ were at increased risk for being COVID-19 positive compared to those with classified as normal weight or overweight. Disentangling the drivers of susceptibility and disease progression will require long-term follow-up in a large, diverse study population, particularly as several comorbidities, such as type 2 diabetes, are also strongly associated with larger body size. Future work from this cohort will include detailed investigations of the impact of body size on susceptibility to and recovery from COVID-19.

Another equivocal risk factor is smoking, which to date has not been clearly demonstrated to convey an increased risk for severe disease (3). In the present work, never smokers comprised 93.5% of the COVID-19 positive participants and 86.1% of the COVID-19 negative participants. This could indicate that those who do not smoke are more susceptible to infection or conversely, that those who smoke are concerned about their risk and are taking additional precautionary measures. A previous study in the United States indicated that current or former smokers were less likely to be hospitalized with COVID-19, but that former smokers were more likely to go on to develop severe disease after hospitalization, and no differences in frequency of critical illness were observed for current smokers(3). However, smoking is known to upregulate the production of the ACE2 receptor cells needed for SARS-CoV-2 to invade cells, though nicotine is known to block the ACE2 receptors(15). This paradox complicates the relationship between smoking and COVID-19. and there is significant variability in the literature. Therefore, more work is needed to assess the role of smoking in COVID-19 disease progression, and future work from CoVHORT will include a detailed analysis of different smoking modalities such as vaping or e-cigarettes, cigar, and cigarette smoking.

Several efforts have been made to identify and characterize the symptom pattern of COVID-19 to allow for more efficient and targeted screening practices, as well as to differentiate SARS-CoV-2 infection from other diseases, such as influenza (8-10, 16). However, these reports of COVID-19 symptoms have largely been confined to observational studies lacking a population-based comparison group. Because many of the symptoms reported as being associated with COVID-19 are general symptoms that could be associated with conditions such as allergies or other infectious illnesses such as influenza, there is an urgent need to evaluate the prevalence of reported symptoms of confirmed COVID-19-positive cases as compared to confirmed COVID-19-negative individuals, as well as with the prevalence of symptoms in the general population.

The results of the present study demonstrate that in southern Arizona, the most common first symptom reported by COVID-19-positive participants was sore throat, other common first symptoms of COVID-19 included headache, cough, runny nose or cold-like symptoms, and fatigue. While these are the same cluster of symptoms as reported by Larsen et al. in a large meta-analysis of more than 50,000 subjects, with data captured by the World Health Organization (WHO), the timing of appearance differed (10). Specifically, the report by Larsen concluded that the order of symptom appearance was estimated to be fever, cough, nausea, and vomiting; while in the current work, the first symptom reported by the majority of cases was sore throat, followed by headache, cough, and runny nose; only 6% of participants had fever as their first symptom. Differences in the study population, including geographic location, sex, age, timing within the pandemic, severity of illness that prompted healthcare seeking behavior and testing, testing accessibility, and race differences across the spectrum of studies employed in the meta-analysis, may explain some of the inconsistent results for first reported symptoms.

An example of this variation in symptom reporting can be observed regarding the number of symptoms that women experienced as compared to men. Women were more likely to be classified in the category of the greatest number of symptoms than men, as were those with a BMI of greater than 30 kg/m^2^, compared to those with a BMI below that threshold, although these findings were not statistically significant. A greater proportion of smokers was observed in the asymptomatic category, as compared to the any symptoms category. These findings suggest that ascertaining the type and order of COVID-19 specific symptomology may be confounded by characteristics of the participants.

With regard to overall symptom profiles, the greatest differences between laboratory-confirmed positive and negative participants were observed for loss of smell and taste and bone or nerve pain, followed by vomiting, nausea, and headache. A similar pattern was seen when comparing cases to the overall untested sample. To date, most work regarding symptoms has relied upon the frequency of symptom occurrence among cases, with little ability to ascertain the degree to which these symptoms differentiate cases from non-cases. For instance, the largest meta-analysis of COVID-19 symptomology to date included data from 24,410 cases from nine countries reported that the most common symptoms were fever (78%), cough (57%), and fatigue (31%) (8). A smaller study within the United States found that the frequency of symptoms among cases was highest for cough (84%), fever (80%), aches and pains (63%), chills (63%), and fatigue (62%)(16). In comparison, herein we found that the most common symptoms reported by cases were fatigue, headache, loss of smell or taste, cough, aches or pains, or sore muscles.

A key finding of this work is that the discrimination of COVID-19-positive symptom profiles from others requires comparison groups. General symptoms reported differ from those which may be applied to differentiate COVID-19 from other infectious diseases or conditions that are present in the underlying population. The symptoms that demonstrated the greatest difference between COVID-19-positive participants and the prevalence of symptoms among laboratory-confirmed COVID-19 negative participants or in the general population were loss of smell and taste, bone or nerve pain, headache, nausea, and fatigue.

The strengths of this study are its prospective nature, ability to capture data for laboratory-confirmed COVID-19-positive cases who have not been hospitalized, and the presence of comparison groups among both those who tested negative for COVID-19 as well as a population base drawn from throughout Arizona. These aspects allowed us to compare symptoms between cases and laboratory-confirmed uninfected individuals. However, limitations of the work must also be considered. Although we have laboratory-confirmed negative participants, we cannot know the COVID-19 status of the untested participants. It is possible that some had already been infected but were asymptomatic or exhibited few symptoms. This would likely attenuate any associations between exposure and outcomes in this study. Additionally, there may be differences in the source population for cases as compared to the laboratory-negative participants and untested participants due to the differences in recruitment strategies for these populations. For example, while postcards were mailed to a random selection of households, it is possible Latinx participants were less likely to respond to this method than direct recruitment as cases during routine case follow-up. This could bias the association between being COVID-19-positive and Latinx away from the null. However, our race/ethnicity profile among cases is approximately similar to the overall distribution of cases throughout Arizona, suggesting a representative sample. Therefore, bias would potentially come from differential responses to other recruitment methods.

In conclusion, the findings of this analysis from the Arizona CoVHORT study show variation in several individual characteristics between COVID-19-positive participants, negative participants, and the untested population, which will be studied in future publications to assess the contributors to these observations. In addition, we found that in southern Arizona, COVID-19 positive participants most commonly reported a sore throat headache, fatigue, cough, or runny nose as the first symptom they noted. These results may aid in earlier identification of cases in the future and highlight the continued importance of addressing surveillance strategies as the pandemic continues.

## Data Availability

Data are not currently available for public use.

